# How to best test suspected cases of COVID-19: an analysis of the diagnostic performance of RT-PCR and alternative molecular methods for the detection of SARS-CoV-2

**DOI:** 10.1101/2021.01.15.21249863

**Authors:** Adrian Mironas, David Jarrom, Evan Campbell, Jennifer Washington, Sabine Ettinger, Ingrid Wilbacher, Gottfried Endel, Hrvoje Vrazic, Susan Myles, Matthew Prettyjohns

## Abstract

As COVID-19 testing is rolled out increasingly widely, the use of a range of alternative testing methods will be beneficial in ensuring testing systems are resilient and adaptable to different clinical and public health scenarios. Here, we compare and discuss the diagnostic performance of a range of different molecular assays designed to detect the presence of SARS-CoV-2 infection in people with suspected COVID-19. Using findings from a systematic review of 103 studies, we categorised COVID-19 molecular assays into 12 different test classes, covering point-of-care tests, various alternative RT-PCR protocols, and alternative methods such as isothermal amplification. We carried out meta-analyses to estimate the diagnostic accuracy and clinical utility of each test class. We also estimated the positive and negative predictive values of all diagnostic test classes across a range of prevalence rates. Using previously validated RT-PCR assays as a reference standard, 11 out of 12 classes showed a summary sensitivity estimate of at least 92% and a specificity estimate of at least 99%. Several diagnostic test classes were estimated to have positive predictive values of 100% throughout the investigated prevalence spectrum, whilst estimated negative predictive values were more variable and sensitive to disease prevalence. We also report the results of clinical utility models that can be used to determine the information gained from a positive and negative test result in each class, and whether each test is more suitable for confirmation or exclusion of disease. Our analysis suggests that several tests exist that are suitable alternatives to standard RT-PCR and we discuss scenarios in which these could be most beneficial, such as where time to test result is critical or, where resources are constrained. However, we also highlight methodological concerns with the design and conduct of many included studies, and also the existence of likely publication bias for some test classes. Our results should be interpreted with these shortcomings in mind. Furthermore, our conclusions on test performance are limited to their use in symptomatic populations: we did not identify sufficient suitable data to allow analysis of testing in asymptomatic populations.

## Background

In December 2019, a novel coronavirus was discovered in Wuhan, Province of Hubei, China and has rapidly spread across the world. This novel coronavirus was named severe acute respiratory syndrome coronavirus 2 (SARS-CoV-2) and causes a disease called coronavirus disease 2019 (COVID-19)^1^. Human-to-human transmission of the virus was confirmed by 30^th^ of January 2020; no population had prior immunity and therefore the entire human population was susceptible to infection2. The World Health Organization (WHO) declared the COVID-19 outbreak a pandemic on the 11^th^ of March 2020.

On the 21^st^ of March 2020, the WHO laboratory testing strategy recommendations highlighted that testing strategies will vary depending on the context of infection status and the testing capacity available in the country in question. The WHO stated that equal access to diagnostic tools is fundamental for limiting the spread of SARS-CoV-2 and adequate testing would contribute to easing of lockdown restrictions and allow societies to reopen3. Several factors can limit testing capacity, including the global availability of kits and reagents4, the turnaround time for test results, and other logistical requirements such as laboratory space and skilled personnel5.

### Current diagnosis guidelines for COVID-19

The WHO recommends that the preferred type of testing for routine confirmation of suspected cases of COVID-19, surveillance testing, contact tracing and environmental monitoring should be based on detection of unique viral RNA sequences by nucleic acid amplification techniques (NAAT), such as RT-PCR^6,7^. The WHO recommends that all suspected cases should be tested in countries that have a limited number of cases, sporadic cases or cluster cases. The objective of this strategy is to rapidly diagnose a high proportion of new cases, and thereby limit the spread of disease via suppressive measures. Nevertheless, in countries with widespread community transmission, testing capacity is often overwhelmed and resources are prioritised for those at greater risk^6^. For example, during the early stages of the pandemic in the United Kingdom, testing was primarily reserved for those who were admitted to hospital^8^.

The laboratory processing of NAAT is complex and the turnaround time for results is approximately 24 hours. Delays and resource dependency can place a strain on testing throughput and capacity. The fine interplay between variables such as the purpose of the testing strategy and capacity should be the main focal points when shaping testing policies and contact tracing programmes in order to re-open economies.

### Alternative molecular tests for the detection of SARS-CoV-2

Molecular tests or commercially available kits are continually being developed in order to increase testing capacity and speed up processing. Alternatives to RT-PCR, such as loop-mediated isothermal amplification (LAMP) or RNA-targeting clustered regularly interspaced short palindromic repeats (CRISPR) have been adapted for rapid and portable detection of nucleic acids. Additionally, some manufacturers have automated some of the processes related to nucleic acid extraction or sample treatment, or even developed automated point-of-care testing (POCT) systems: these are often cartridge-based provide test results via a much simpler process and in a shorter time than laboratory-based RT-PCR.

These variations in procedures and methods reflect the wide range of SARS-CoV-2 tests that have proliferated since the beginning of the COVID-19 pandemic. Each variation will have its own advantages and may be utilised depending on the purpose of testing and the context in which it is carried out. For example, skipping the extraction process could increase the capacity of a testing programme due to shorter turnaround times and the avoidance of extra costs associated with extraction kits. Furthermore, POCT systems can alleviate the need for specialised laboratory personnel. It is therefore important to evaluate the impact of different tests and methods to facilitate informed decisions when planning a public health testing strategy. To our knowledge, no research has previously been carried out to systematically evaluate and compare the diagnostic performance of different NAAT methods and protocols.

### The role of rapid collaborative reviews in the COVID-19 response

During the COVID-19 pandemic, timely and reliable assessment of evidence helps develop a co-ordinated response and inform both healthcare professionals and the general public. The European network for Health Technology Assessment (EUnetHTA) spans more than 80 Health Technology Assessment (HTA) partners across Europe. The mission of EUnetHTA is to support collaboration between European HTA organisations that brings added value to healthcare systems at the European, national, and regional level, making it well-positioned to respond to the COVID-19 pandemic^9^. The primary objectives of EUnetHTA during the COVID-19 pandemic are to provide decision-makers with timely syntheses of available evidence on the safety and effectiveness of health technologies for the management of the current pandemic, as well as providing timely updates as research evolves and the relevant body of evidence matures^9^.

Following the EUnetHTA Plenary Assembly in April 2020, a EUnetHTA Task Force on SARS-CoV-2 diagnostics was set up, which prioritised the following health policy questions:

1. How to best screen asymptomatic subjects and monitor close contacts in order to promptly detect infections among the general population and healthcare workers.
2. How to best test patients with clinical manifestations of SARS-CoV-2 in order to confirm the diagnosis of COVID-19.
3. Which tests should be used to monitor the course of disease and inform decisions on treatment, hospitalisation etc. and to determine viral clearance of recovered patients in order to allow re-entry into the community.

EUnetHTA has prioritised COVID-19-related outputs until the end of May 2021, producing ‘Rapid Collaborative Reviews’ (RCROT) for diagnostic testing, ‘Rapid Collaborative Reviews’ (PTRCR) and ‘Rolling Collaborative Reviews’ (RCR) for therapeutic treatments. Here, we summarise the findings and policy implications of the second RCROT, on the diagnostic accuracy of molecular methods that detect the presence of the SARS-CoV-2 virus in people with suspected COVID-19. A previous RCROT on the role of antibody testing was published in June 202010. This work was part of the project undertaken by the EUnetHTA Task Force on SARS-CoV-2 diagnostics and it addressed the policy priority questions (1) and (2).

### The scope and purpose of the rapid collaborative review

As COVID-19 testing continues to be scaled up and used more widely, there is a clear need to evaluate alternative molecular methods and approaches to allow NAAT to continue in the face of potential challenges and global shortages of kits and reagents, and to inform decisions about where different test methods and protocols are best deployed. Our evaluation of the diagnostic accuracy of different molecular tests and methods aimed to address the questions above, but also to determine the diagnostic performance of different test classes. This could facilitate the identification of novel alternative tests that could assist with, for example, a wider rollout of COVID-19 testing or tests that can be used reliably where a rapid result is needed.

The full scope of the assessment has been previously published in the project plan^11^. Briefly, we carried out a systematic review and rapid HTA that aimed to address the following research question: What is the diagnostic accuracy of molecular methods that detect the presence of the SARS-CoV-2 virus in people with suspected COVID-19? We included studies of any population tested for COVID-19 (whether for diagnosis on the basis of clinical symptoms, contact tracing or as part of mass screening), which studied any molecular assay based on nucleic acid amplification to detect the presence of SARS-CoV-2 infection, and compared the studied test(s) against a suitable reference standard (RT-PCR conducted on specific targets of SARS-CoV-2 virus using a validated assay, alone or in combination with clinical findings, as per the WHO recommendations). We only included studies if they reported the numbers of true/false positive participants and true/false negative participants, and used this to calculate diagnostic accuracy outcomes both for individual studies or pool diagnostic accuracy outcomes across studies, stratified according to the characteristics of each index test. Full details of study assessment, data analysis and synthesis have been reported elsewhere^12^.

### Description of the evaluated evidence

Our analysis of diagnostic accuracy was based on data from 103 primary studies that met the inclusion criteria for this review (62 articles published in peer-reviewed journals and 41 pre-prints). In 33 studies, multiple tests were studied: there was variation in protocols/kits/platforms, methods of nucleic acid extraction and processing, targets, sample types, or comparison of the same test to more than one reference standard. This meant a total of 168 observations (2×2 contingency tables, reporting numbers of true/false positive participants and true/false negative participants for each test) could be extracted from the primary studies. From those, 18 observations were excluded, either because they were conducted on populations not relevant to this review, or in order to avoid data duplication within the pooled analysis. Originally, we planned to analyse test performance in symptomatic populations, and also use in asymptomatic populations as part of mass screening or contact tracing. However, we found very little evidence on asymptomatic or convalescent populations that met our inclusion criteria, and therefore our analysis focusses on use of tests in symptomatic populations.

We classified the evidence found into 12 distinct diagnostic test classes based on the shared commonalities of the index test assessed in the context of that study. Table 1 describes each diagnostic test class, and the number of studies and observations for which data was available for each class.

**Table 1.** Delineated diagnostic test classes with associated descriptions, number of studies and observations.

| **Index Test Class** | **Class Description** | **Number of Studies** | **Number of Observations** |
| --- | --- | --- | --- |
| **Automated RT-PCR Systems** | Integrated, high throughput, fully automated laboratory workflow systems | 10 studies | 10 observations |
| **Commercial RT-PCR Kits** | Manual commercial reagent kits based on RT-PCR technology | 13 studies | 25 observations |
| **POCT Systems** | Automated, rapid point of care testing based on cartridge technologies | 20 studies | 29 observations |
| **Different RT-PCR Methods** | In-house, laboratory derived assays with variations in the assay technique and method, based on RT-PCR technology | 16 studies | 23 observations |
| **RT-qPCR** | Manual laboratory assays based on quantitative RT-PCR technology | 6 studies | 9 observations |
| **RT-RAA** | Manual laboratory assays based on reverse transcriptase recombinase-aided amplification technology | 4 studies | 4 observations |
| **RT-nPCR** | Manual laboratory assays based on reverse transcriptase nested PCR technology | 3 studies | 4 observations |
| **dRT-PCR** | Manual assays based on digital RT-PCR technology | 3 studies | 3 observations |
| **LAMP** | Manual assays based on LAMP technology with colorimetric, automated or naked eye detection | 8 studies | 8 observations |
| **RT-LAMP** | Manual assays based on RT-LAMP technology with multiple variations in methods and colorimetric, automated or naked eye detection | 20 studies | 24 observations |
| **TMA** | Manual laboratory assay based on TMA technology | 4 studies | 4 observations |
| **CRISPR** | Manual laboratory assay based on CRISPR technologies | 6 studies | 7 observations |
| CRISPR: clustered regularly interspaced short palindromic repeats, dRT-PCR: digital reverse transcriptase polymerase chain reaction LAMP: loop-mediated isothermal amplification, POCT: point-of-care testing, RT-PCR: reverse transcriptase polymerase chain reaction, RT-nPCR: reverse transcriptase nested polymerase chain reaction, RT-qPCR: reverse transcriptase quantitative polymerase chain reaction, RT-RAA: reverse transcriptase recombinase aided amplification, RT-LAMP: reverse transcriptase loop-mediated isothermal amplification, TMA: transcription-mediated amplification |  |  |  |

### Pooled analyses and clinical utility models

We carried out pooled analyses for each diagnostic test class to evaluate their diagnostic accuracy and clinical utility. Diagnostic accuracy estimates were generally high and similar across classes: 11 out of 12 classes showed a summary sensitivity estimate of at least 92% and a specificity estimate of at least 99%. The LAMP test class had the lowest summary estimate for sensitivity (87%) whilst the minimum estimate for specificity was for dRT-PCR (83%). The area under the curve (AUC) values of the summary receiver operating characteristics (ROCs) showed a minimum value of 0.98, which is far greater than the reference value of 0.70 for risk prediction in diagnostic tests^13^.

Clinical utility modelling showed greater variability across the 12 diagnostic test classes. The probability modifying plots for test classes such as Automated RT-PCR Systems, RT-qPCR and RT-RAA suggest that positive and negative test results are equally informative. The plots for five other classes indicated that the tests have marginally more informative positive results than negative. Commercial RT-PCR Kits, LAMP and RT-LAMP are the most skewed classes towards informative positive results. Likelihood ratio scattergrams indicated that, with the exception of LAMP, all the evaluated test classes were likely to be useful for both confirmation and exclusion of disease. Similarly, Fagan plots showed that for patients who were under suspicion of infection, the probability of having the disease was reduced to 0% when the test result was negative for all tests classes, with the exception of LAMP. With regard to positive post-test probability, only the RT-nPCR class showed a maximum value of 100%, indicating that in patients who are suspected of infection, the probability of having the disease was 100% after a positive test. The second highest post-test probability was showed by the Commercial RT-PCR Kits and other test classes such as Automated RT-PCR Systems, Different RT-PCR Methods, POCT Systems, LAMP, TMA and CRISPR had a positive post-test probability of at least 90%. Table 2 summarises the pooled estimates of sensitivity and specificity and the results of the different clinical utility models for the 12 different diagnostic test classes.

**Table 2.**
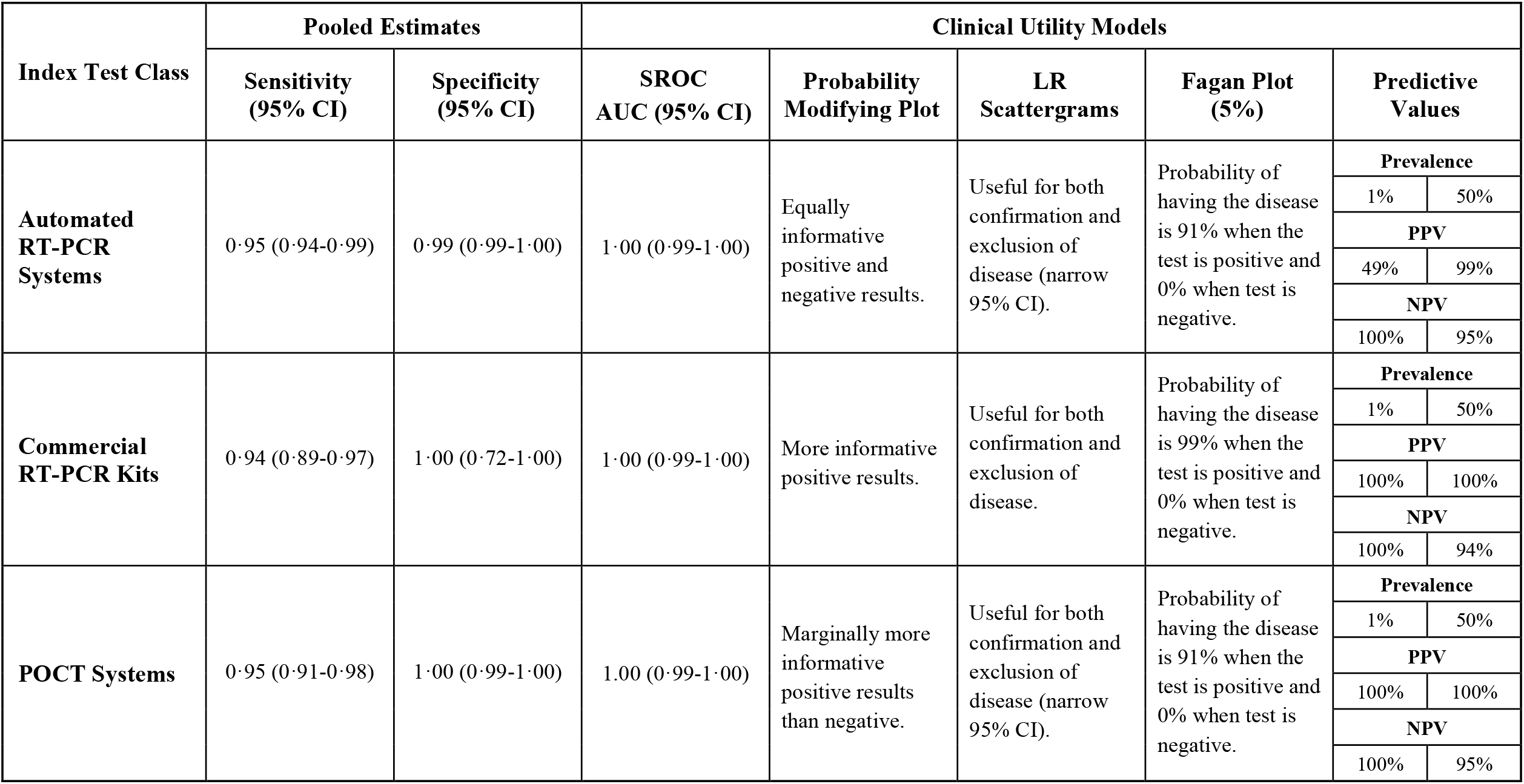
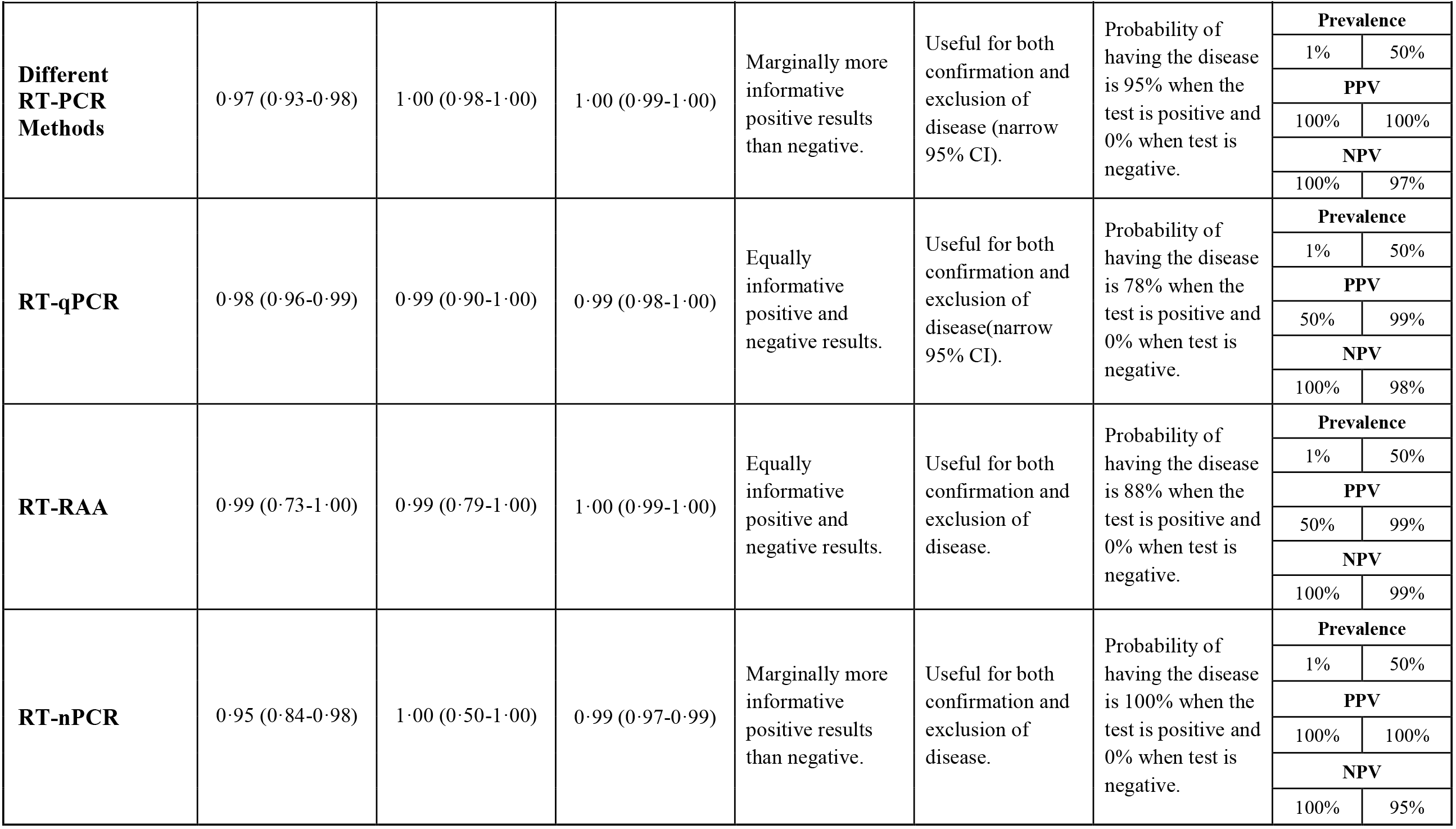
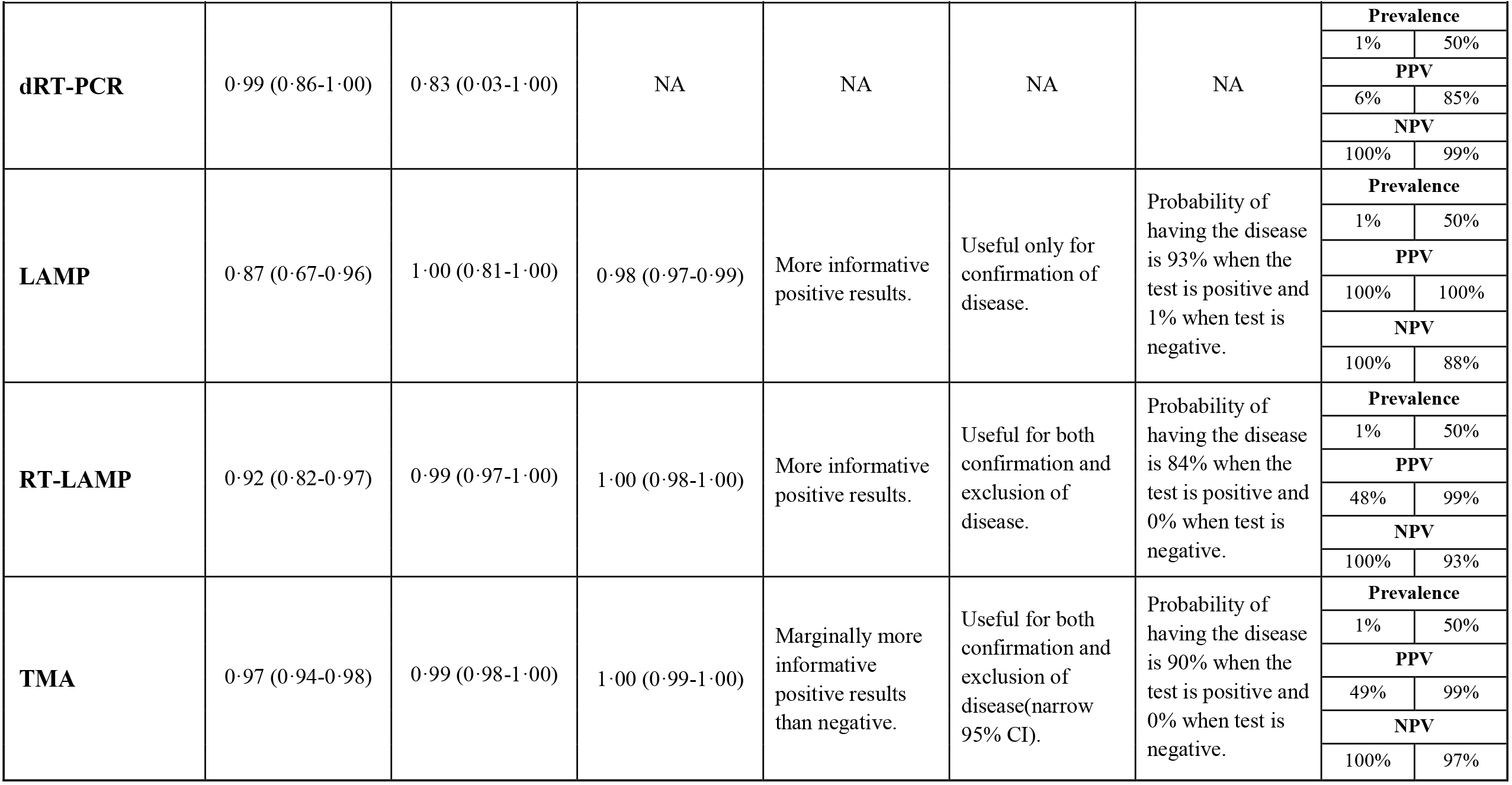
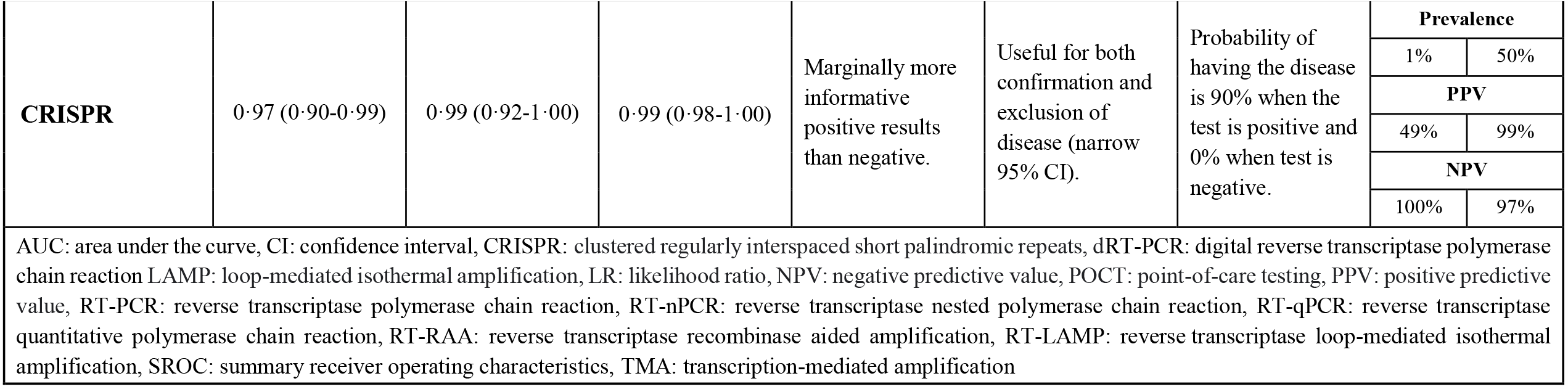
Summary of diagnostic accuracy estimates and performance of the diagnostic test classes as reflected by the pooled estimates and clinical utility models.

In order to facilitate the translation of the results into clinical settings, we estimated the positive and negative predictive values of all diagnostic test classes across a range of prevalence rates. Certain diagnostic test classes were found to retain positive predictive values (PPV) of 100% throughout the investigated prevalence spectrum. These technologies include Commercial RT-PCR Kits, POCT Systems, Different RT-PCR Methods, RT-nPCR and LAMP. However, there is variation in the negative predictive values (NPV) across the prevalence spectrum, with Commercial RT-PCR Kits, POCT Systems, Different RT-PCR Methods and RT-nPCR retaining a minimum value of 94% at the highest threshold of 50% prevalence. A graphical representation of the PPV and NPV across the prevalence rates for the test classes can be found in Figure 1 and 2.

**Figure 1.**
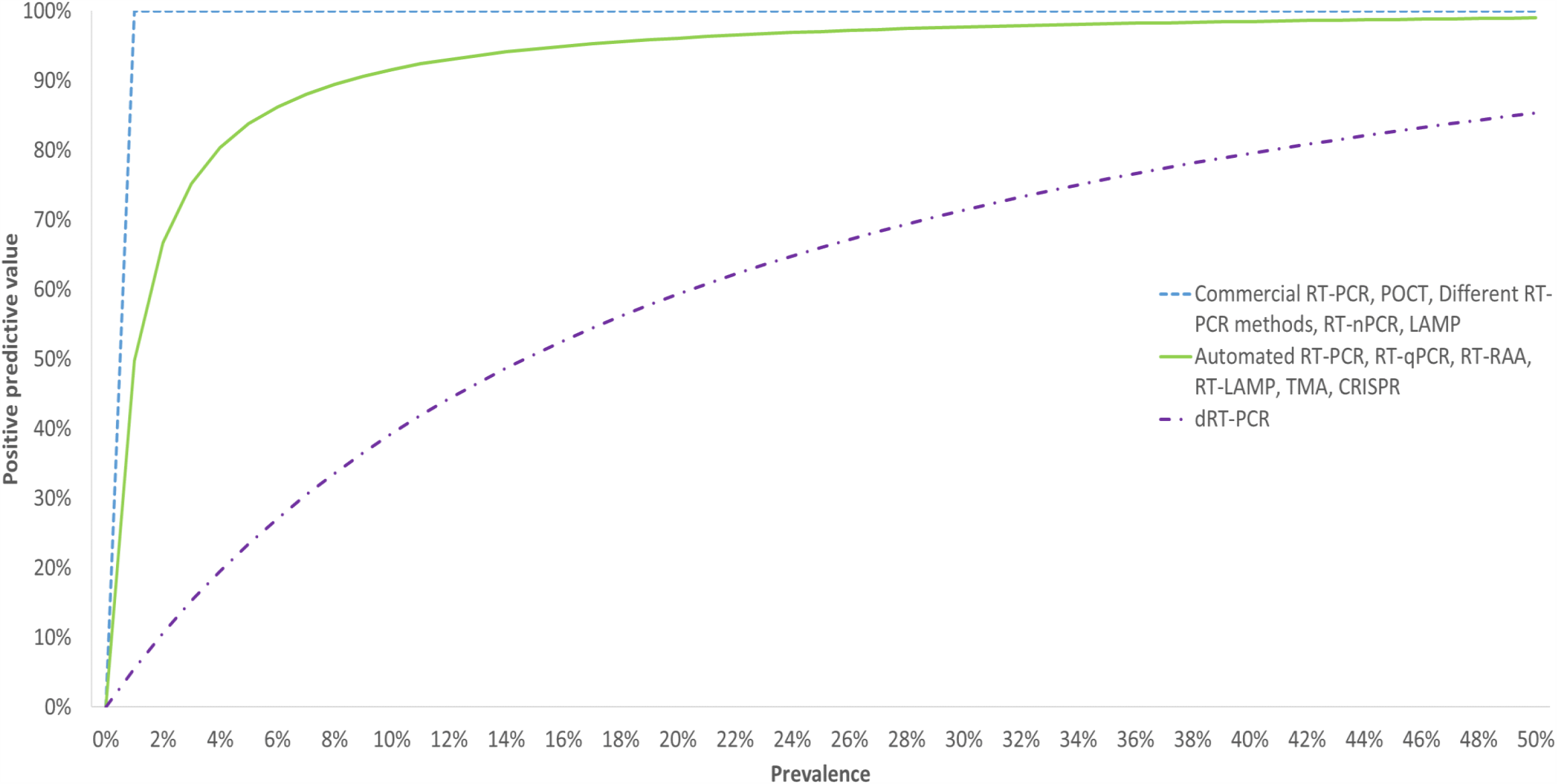
Positive predictive values for the test categories across a range of prevalence rates.

**Figure 2.**
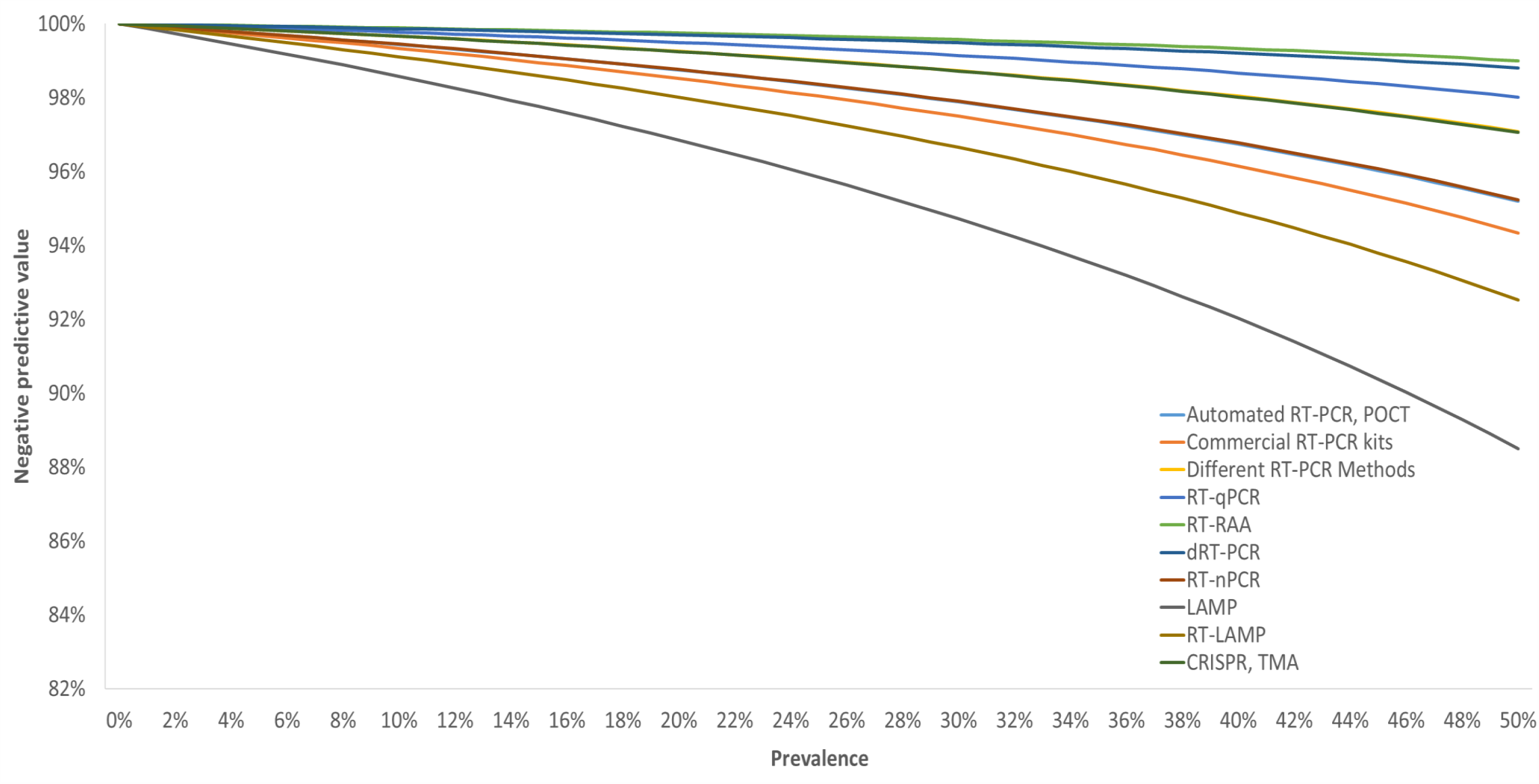
Negative predictive values for the test categories across a range of prevalence rates.

### Evaluation of the methodological quality of the included studies and publication bias

The quality of the extracted observations was assessed using the Quality Assessment of Diagnostic Accuracy Studies 2 (QUADAS-2) tool^14^. The patient selection domain had the most observations at high or unclear risk of bias (70%). The majority of the included studies in this review followed a case-control design (59/103), which in this context means the population would have consisted of some subjects known to have COVID-19 (‘cases’) and some subjects known to be free of disease (‘controls’). Studies of this design less closely reflect the real-world application of these tests and increase the likelihood of variation of the test performance in a clinical setting. Similarly, in the index test domain, 64% of the observations were classified as a high or unclear risk of bias. As with patient selection, the classification was based on concerns originating from study designs where the index test was not carried out without prior knowledge of the disease status or reference standard test result. In comparison, the majority of observations had clear reporting of the reference standard, used an appropriate time interval between the index test and reference standard being carried out and clearly accounted for all recruited patients in their reported results. We therefore classed most studies as low risk of bias in the reference standard (82%) and flow and timing domains (65%). Low applicability concerns were generally reflected across the patient selection (93%), index test (97%) and reference standard (98%). Publication bias was assessed for each of the delineated diagnostic test classes using the Deeks’ funnel plot asymmetry. A significant publication bias was found in the following classes: POCT Systems, Different RT-PCR Methods, RT-qPCR and CRISPR. This suggests that studies with positive results are more likely to be published for those classes than studies indicating more negative results.

### Methodological limitations and performance considerations

Although the evaluated diagnostic test classes show comparable accuracies across the spectrum, certain factors could reduce the generalisability of the findings. The lack of data reflecting tests conducted in asymptomatic and convalescent patients or as part of contact-tracing programmes as well as the selection of symptomatic cohorts may lead to the introduction of bias that in turn could produce overestimates of the test accuracies. Another important limitation to acknowledge is related to the categorisation of the observation into the 12 diagnostic test classes. We aimed to classify tests in a way that allowed results from similar studies to be pooled, without inappropriately grouping together heterogeneous data. As there is no standardised classification system for tests that detect SARS-CoV-2 using molecular methods, we derived our own suitable classification system and associated definitions for these. Although we attempted to closely match the observations and divide them into classes based on their shared commonalities, inevitably there will remain some differences between tests grouped within the same class, either in terms of their known characteristics or because of factors that were not reported in the included studies. For studies that reported more than one observation and it was not clear if the patient population was different, where possible we attempted to avoid any data duplication by excluding those observations from the analysis. Nevertheless, we could not completely rule out the possibility of data duplication in all cases and therefore this is another potential limitation of our analysis.

Although the assessment initially aimed to address two policy priority questions established by the SARS-CoV-2 EUnetHTA Task Force, it was not possible to evaluate the accuracy of NAAT to screen asymptomatic subjects or monitor close contacts due to the lack of evidence evaluating molecular tests and methods in those populations. We believe that this is an important observation at this stage in the global pandemic and we suggest that rollout of mass screening programmes should be used as an opportunity to conduct more high-quality diagnostic accuracy studies in those populations, particularly where they use alternatives to standard RT-PCR-based testing. Studies should also aim to report the time since infection in order to ascertain the diagnostic accuracy and performance of molecular tests for monitoring infectivity and suppressing transmissibility. Furthermore, future research in this field should aim to report complete and uniform data sets, containing all variables necessary for future explorations of effect modifiers and confounding factors of the diagnostic performance of different tests or methods (e.g. patient characteristics, demographics, symptoms, etc.).

### Testing policy implications

As aforementioned, we generally found different evaluated test classes to have comparable diagnostic accuracies and performances, as indicated by our clinical utility models. The stratified test classes reflect different molecular tests that are fundamentally diverse in regards to the underlying technologies, methods applied or detection principles. Therefore, it is essential to highlight the potential advantages and disadvantages of using different molecular methods for the detection of SARS-CoV-2 in order to inform policy and decision makers about viable alternatives for the COVID-19 testing strategies. The WHO recommendations indicate that testing strategies should be developed for the situational context and not vice-versa^6^. Each testing modality may provide advantages in different situations. Advantages of the molecular testing strategies can be broadly grouped into three categories: time to test result, cost, and other resource requirements.

### Implications of time to test result

Hospital emergency departments exemplify a situation where both time to result and a high diagnostic test accuracy for the diagnosis of COVID-19 is desired. For example, the ability to have a highly accurate and rapid turnaround for test results would allow decisions to be taken on whether to isolate a critically ill patient as soon as possible after admission. Our analysis suggests POCT Systems are both highly accurate and rapid in their turnaround time. From a different perspective, frontline staff such as healthcare and other key workers could also highly benefit from diagnostic platforms that produce quick results in a highly accurate manner. This would allow a prompt initiation of isolation of the frontline workers, suppression of further transmission of the virus and would decrease the strain placed on the health services which are dealing with personnel that is in isolation. Similarly, long term care facilities for the elderly could also benefit from testing strategies embedding POCT Systems as they could allow the rapid isolation of both staff and residents should they became symptomatic. Furthermore, settings that provide care for the elderly could also benefit from such testing strategies since this patient group has been highlighted of being both at high risk and having a high mortality rate in the pandemic^15^.

Airports and other transit hubs represent other settings that could benefit from a short turnaround time for diagnostic tests. Currently, the majority of countries have very limited travel corridors and will not accept incoming passengers from countries classified at high risk of infection. The ability to test passengers on arrival could help bypass the need for such travel corridors or to ease restrictive measures such as the need for pre-travel quarantine. The ability to accurately and rapidly test individuals could remove existing barriers to international travel and help reopen the economy for both commerce and tourism. However, this would represent use of tests for mass screening, a scenario for which we found very little evidence compared to that available for symptomatic testing, and this is therefore an area where more research is required.

Automated RT-PCR Systems use high throughput platforms and have the ability to process hundreds of samples simultaneously, increasing testing capacity while retaining diagnostic accuracy, with the same amount of skilled personnel and laboratory space. Notably, some of the systems have automated the entire testing process (lysis, extraction, PCR). Traditionally, fully automated RT-PCR Systems have been considered to be less sensitive than conventional RT-PCR testing protocols^16^. However, this review found that accuracy and performance of Automated RT-PCR Systems was comparable to manual RT-PCR approaches and had a higher sensitivity than Commercial RT-PCR Kits (Table 2). Using such platforms to boost testing capacity could allow some countries to either initiate or consolidate the bases of test and trace programmes. For example, in the United Kingdom, a ‘test, trace, isolate’ system is used. However, if the testing capacity was expanded it could allow the implementation of models to facilitate the reopening of the economy by decreasing the number of people who need to isolate as a precaution whilst awaiting a test result.

### Implications of cost and resource requirements

A lack of testing or reduced testing capacities have been highlighted as significant issues in low- and middle- income countries^17^. Additionally, the complete or reduced activity of the economies in low- and middle-income countries has also been acknowledged to have more severe effects on poverty and health inequalities^18^. In these situations, the application of less costly and resource-intensive molecular tests such as RT-LAMP and CRISPR could provide alternatives with comparable diagnostic performances to RT-PCR. This implies that countries which currently face both resource and financial challenges for the establishment of a comprehensive testing program could potentially use these alternative methods and still retain a high level of diagnostic accuracy. Furthermore, the simplification of protocols by performing some molecular tests on samples without requiring a nucleic acid extraction step has been shown to have a marginal effect on the diagnostic accuracy of these tests^12^. This could alleviate the global shortages of extraction kits and could present an opportunity to establish testing programmes which may not reflect the highest performance, but could be used with a degree of confidence that diagnostic accuracy should be retained when testing symptomatic individuals.

### Final considerations

We urge policy makers to interpret the findings of this review with caution and to consider all the strengths and limitations when designing testing strategies based on different molecular tests. It is essential to acknowledge that clinical relevance and performance of different diagnostic tests are highly dependent on the disease prevalence rates. The WHO urged research for the development and evaluation of simpler and more portable detection platforms for a reliable diagnosis of COVID-19 in order to suppress transmission, identify close contact, understand the disease epidemiology, monitor responsiveness to treatment and the impact of public health and social measures^7^. These alternative diagnostic test classes are reliant on different technologies that could provide access to testing in locations with limited laboratory capacity and have a rapid turnaround time for the generation of the results. Lastly, we re-emphasis the need for future research on the evaluation of diagnostic accuracy of all test methods in asymptomatic and convalescent populations or as part of contact screening programmes.

## Supplementary material

The rapid collaborative review, project plan and plain language summary can be found at: https://eunethta.eu/rcrot02/

## Search strategy and selection criteria

We searched Ovid Medline, Ovid Embase, Cochrane Central Register of Controlled Trials (CENTRAL), Cochrane COVID-19 Study Registry, ClinicalTrials.gov, WHO International Clinical Trials Registry Platform, EU Clinical Trials Registry, pre-print servers and pre-defined key websites. The following key concepts (and their synonyms) were used to develop a comprehensive search strategy for articles published from 1^st^ January to 29^th^ July 2020: SARS-CoV-2; COVID-19; Molecular Tests; Nucleic Acid Amplification Test; Reverse Transcriptase Polymerase Chain Reaction & Reverse Transcriptase Loop-Mediated Isothermal Amplification. References were included based on their relevance to the broad scope of this review.

## Authors’ contribution

Adrian Mironas (AM) has contributed to the conceptualisation, methodology, investigation, data collection and curation, formal data analysis and interpretation, visualization, writing of original draft and review editing in addition to the verification/validation of the underlying data extracted by the rest of the authors.

David Jarrom (DJ) has contributed to the conceptualisation, investigation, technical oversight, supervision methodology, validation, writing of original draft and review editing.

Evan Campbell (EC) has contributed to the conceptualisation, investigation, data collection and curation, risk of bias analysis, validation and review editing.

Jennifer Washington (JW) has contributed with the literature search, created the methodology for information retrieval and resources associated with literature retrieval and formatting.

Sabine Ettinger (SE) has contributed with the project administration.

Ingrid Wilbacher (IW) has contributed to the conceptualisation, investigation, data collection and curation, risk of bias analysis and validation.

Gottfried Endel (GE) has contributed to the conceptualisation, investigation, data collection and curation, risk of bias analysis and validation.

Hrvoje Vrazic (HV) has contributed to the conceptualisation, investigation, data collection and curation, risk of bias analysis and validation.

Susan Myles (SM) has contributed with conceptualisation, investigation, methodology, supervision, technical oversight and validation.

Matthew Prettyjohns (MP) has contributed with conceptualisation, investigation, methodology supervision, technical oversight, formal data analysis, validation and visualisation.

## Declaration of interests

The rapid collaborative review and the resulting manuscript were established within the European Network for Health Technology Assessment (EUnetHTA), which received funding from the European Union’s Health Programme (2014-2020) for the project / joint action ‘724130 / EUnetHTA JA3’. AM, DJ, MP, JW, SM received non-financial support from EUnetHTA due to their Associate Membership, the remainder of authors as EUnetHTA members received co-funding.

The content of this publication represents the views of the author(s) only and is his/her sole responsibility; it cannot be considered to reflect the views of the European Commission and/or the Consumers, Health, Agriculture and Food Executive Agency or any other body of the European Union. The European Commission and the Agency do not accept any responsibility for use that may be made of the information it contains.

## Data Availability

The rapid collaborative review, project plan and plain language summary can be found at the link provided below.

https://eunethta.eu/rcrot02/

## Acknowledgements

We would like to extend our gratitude to the EUnetHTA RCROT02 assessment team that was involved in the rapid collaborative review, in particular those who reviewed and commented on the draft review and on the project plan.

